# The determinants of malaria prevention practices in the pastoral communities of the South Omo zone, Ethiopia

**DOI:** 10.1101/2025.05.28.25328516

**Authors:** Abrham Mogga Gimaro, Pattrone Rebecca Risenga

## Abstract

Malaria, a global health problem, primarily affects people in developing countries in tropical and subtropical regions, causing severe illness and death if not treated promptly. Despite being a preventable disease, it continues to pose a significant threat to communities, such as pastoralists. As a result, this study assessed the factors that influence malaria prevention practices pastoralists in the South Omo zone, Ethiopia.

A cross-sectional community-based study was conducted in the South Omo Zone in Ethiopia. A face-to-face questionnaire was used to collect data from 653 pastoral households living within five high-malaria woredas. A multistage cluster sampling technique with systematic sampling was used to select the households from different clusters. The data was analysed using SPSS version 28 for descriptive statistics, binary, and multivariable logistic regression.

Multivariable logistic regression revealed that the odds of practicing at least two and possibly more malaria prevention measures were significantly higher in those with long-lasting insecticide-treated nets at home (adjusted odds ratio (AOR) = 7.881, 95% confidence interval (CI) = 2.309– 26.896), those who understood the concept of malaria prevention (AOR = 3.399, 95% CI = 2.106– 4.915), female-headed households (AOR = 1.616, 95% CI = 1.115–2.334), and middle-income households (AOR = 2.746, 95% CI = 1.534–4.915) were more likely to practice at least two and possibly more malaria prevention measures in the wealth index quantiles.

Socioeconomic status, access to preventive measures, awareness of malaria prevention and households structure play significant roles in the likelihood of practicing a combination of malaria prevention measures.

Therefore, efforts to reduce malaria transmission have to prioritize improving access to preventive measures and malaria prevention education, and interventions addressing socioeconomic disparities among pastoral households may aid in preventive measure adoption.

## Introduction

Malaria is a global health problem that predominantly affects people of developing countries living in tropical and subtropical areas [1].

Globally, there were 263 million estimated malaria cases and 597,000 malaria-attributed deaths that occurred in the year 2023 [2]. The WHO African region accounts for 95% of the reported malaria cases in 2023, and the majority of malaria cases are Plasmodium falciparum (P.F.) [2, 3]. Asia is the second malaria-endemic continent after Africa, covering 3.4% of the global malaria burden [4], and malaria is endemic in 20 countries around South and Southeast Asia, where India accounts for three-fourths of the total malaria burden of Southeast Asia [3, 4]. In Latin America and the Caribbean, there are 21 malaria-endemic countries, and Plasmodium vivax (P.V.) is the dominant malaria parasite, accounting for 72% of the total malaria cases on the continent [5: P57].

In Ethiopia, in the year 2023; malaria caused an estimated 7.3 million new cases and 1,157 deaths in the country, the most cases in a single year in the previous seven years [6, 7]. The two dominant malaria parasite species; *Plasmodium* falciparum and *Plasmodium* vivax, are accountable for 60% and 40% of the malaria infections in the country, respectively [8].

Malaria is a significant challenge to social and economic development in Ethiopia, and it is prevalent in 68% of the country’s landmass [9]. According to the World Malaria Report 2024, Ethiopia has one of Africa’s highest malaria burdens, accounting for 4% of all malaria cases in 2023 [2].

Every year, malaria is responsible for an estimated 5 million cases, 70,000 deaths, 17% of outpatient visits, 15% of admissions, and 29% of inpatient deaths in the country [10]. The incidence and prevalence of malaria rise during the planting and harvesting seasons. According to different studies conducted in the country, the prevalence of malaria during planting season extends to 18.4% among mobile labour workers [11, 12], while its prevalence is below 1% in non-planting and non-harvesting seasons [13].

Malaria disproportionately affects vulnerable groups like *nomadic pastoralists*, migrants, refugees and displaced people due to their mobility and inaccessibility to health service provision [14]. The disease has been occurring in a form of epidemics and results in a high mortality among nomadic and seminomadic pastoralists due to their mobility, difficulty in access and live-stock centeredness [15, 16]. As a result, they deserve extra malaria prevention efforts in addition to pregnant women, under 5 children and visitors from non-endemic countries [17].

Pastoralists are defined as groups of people who rely primarily on the products of their hoofed domestic animals and tailor their settlement and mobility strategies to their livestock’s dietary needs, often moving seasonally to find fresh grazing land [18].

Dong and Kassim define pastoralism “as a production system and livelihood strategy based on extensive livestock grazing on rangelands/grasslands, often with some form of herd mobility, that has been practiced in many parts of the world for centuries” [19]. Pastoralism is a way of life that is deeply rooted in traditional knowledge and practices, enabling communities to adapt to harsh environmental conditions while also maintaining long-term relationships with their natural surroundings [20, 21].

Globally, the numbers of pastoralists account for approximately 50-100 million peoples [22]. In Africa, pastoralism as a way of life covers arid and semiarid land from Mauritania to Senegal in west and parts of Ethiopia, Kenya and Somalia in East [23] and Africa serves as the home for more than 60% of the nomadic pastoralists [24].

In Ethiopia, pastoralists generally live in the drylands of 4 regional states, namely, Afar, Somalia, Oromia (Borena zone), and Southwest and South Ethiopia region’s (South Omo zone, where the study area is located) [25], and they nationally account for about 10 million people [26]; and the terminology preferably used in Ethiopia nowadays is pastoralists instead of nomads. Despite the existence of some estimates at the global level, it is difficult to have the correct estimates of pastoralists living elsewhere due to their mobility and limited accessibility.

Since the year 2005, Ethiopia has been implementing rapid scale-up of the four proven antimalarial interventions: insecticide-treated mosquito nets, indoor residual spraying, correct diagnosis, and early treatment with artemisinin-based combination therapies [27]. This has resulted in a marked reduction in the incidence and prevalence of malaria in the country. The example of this reduction was that the number of estimated new malaria cases that occurred declined from 2.8 million in 1990 to 621,345 in 2015 [28].

Despite the application of malaria control and preventive measures and remarkable achievements recorded in most parts of Ethiopia, an increment in the incidence and prevalence of malaria in the South Omo administrative zone has been reported. According to the 2018 annual malaria control and elimination report of the Southern Nations, Nationalities, and Peoples’ Region (SNNPR), 30% of the SNNPR’s malaria burden was reported from the South Omo administrative zone, while the population share of this zone constitutes 4% of the former Southern Nations, Nationalities, and Peoples’ Region (SNNPR) population [29]. A study conducted in the Benatsemay district of the South Omo zone revealed that the average attack rate of 114/1000 population, which is higher than the usual malaria incidence, was observed in most of the other districts [30].

As the area is remote and difficult to access, there was a shortage of published literature on malaria prevention control interventions highlighting the need for further research and targeted interventions to address the high malaria burden in South Omo.

Because the South Omo zone is remote and difficult to access, little research has been published on the pastoral community’s methods for preventing and controlling malaria, as well as seeking treatment.

As a result, the aim of this study was to identify factors influencing malaria prevention practices in the South Omo zone in Ethiopia.

## Materials and methods

### Study design and study setting

A cross-sectional community-based study was conducted between 18^th^ April 2021 and 10^th^ June 2022 in five purposively selected high-malaria-burden districts of the South Omo administrative zone, Ethiopia. These five districts are Salamago, Benatsemay, Hamer, Malie, and Daasenech.

The study included pastoral household heads from five high malaria burden districts in the South Omo zone, with ages ranging from 18 to 70, who were willing to participate and had lived in the area for at least a year. It did, however, exclude household heads who were deaf, blind, or seriously ill, as well as those who refused to participate and had only lived in the area for one year.

The South Omo administrative zone is one of 12 administrative zones in South Ethiopia region. The South Omo zone got its name after the Omo River, which flows south into Lake Turkana on the western coast of Kenya [31]. The South Omo administrative zone is located about 750 kilometres south of Addis Ababa, the capital city of Ethiopia. Dimeka serves as the administrative centre for South Omo.

This zone’s administrative divisions consist of eight districts and two town administration. The South Omo administrative zone is bordered by Kenya to the south, West Omo Zone to the west, Keffa Zone to the Northwest, Ari Zone and Gofa Zone to the North, Gardula, Ale Zone, and Konso to the Northeast, and the Oromia Region to the East. Animal husbandry is a major economic activity in the South Omo administrative zone, with cattle, goats, and sheep being the most commonly raised animals [32].

Mago National Park and Tama Wildlife Reserve are located on the Omo River’s eastern bank [33]. This zone, which is home to pastoral ethnic groups, is one of the country’s least populous areas. The population of the South Omo administrative zone was estimated to be 447,726 in the year 2025 [34]. The territory is characterized by poor or non-existent infrastructure and challenging terrain access [33].

### Operational definition

#### Long-lasting insecticide nets (LLINs) use

The data collector observed a standardized, properly mounted (hung) net used over the bed and sleeping space that was free of holes or tears, as well as household members sleeping with their LLINs the night before the survey.

#### Malaria prevention practices

Malaria prevention was considered “practiced” when the head of the household had used at least two of the three measures (long-lasting insecticide-treated nets, indoor residual spraying, and/or larvicide) in the 12 months prior to this study. This construct, or composite measure, was determined by computing the response of the head of the household as an average score of the sum of all measures used, ranging from 0 to 3 and greater than or equal to 2 was considered to be practiced.

#### Indoor residual spray

The housing structure was considered an *“indoor residual spray* (IRS)” when the internal walls and ceilings of housing structures or nomadic camps were sprayed with residual insecticide chemical within the last 12 months preceding the data collection and responded “yes” to the question, “Has your house been sprayed with insecticide in the last 12 months?”

**Attitude towards malaria prevention:** was measured by computing the total score of responses given by the participant to the 7 attitude statements with the 5-point Likert scale and then comparing it against the attitude mean score, which was 28.38. Participant’s score ≥ mean score (28.38), was categorized as a “*good attitude” or and a* participant’s *score below the mean was* labelled as a “poor attitude.

#### Knowledge of malaria prevention

Respondents’ knowledge of malaria prevention was measured by calculating the mean score of correct responses to the 4 knowledge related questions. The knowledge related questions were about what causes malaria, mosquito preventive measures like IRS application and utilization of LLINS and dreaming of stagnant water. Participants with a correct mean score of 1.75 or above were labelled as “knowledgeable,” while those with a score below the mean but not all of the correctly answered questions were labelled as “not knowledgeable.”

#### Wealth index

The wealth index, which is the relative economic situation of pastoral households, was measured using 12 variables like housing, livestock, agricultural land, household services, and bank accounts. Principal component analysis is applied to construct a wealth index, ranking wealth into five quantiles: extremely poor, poor, middle class, wealthy, and extremely wealthy, based on housing conditions and non-productive assets.

#### Data source and sampling

Face-to-face interviews were conducted with pastoralist households living in five high-malaria-burden woredas (Malie, Hamer, Benna Tsemay, Nyangatom, and Selamango) in the South Omo administrative zone, Ethiopia. The study included 22 clusters from these five malaria-prone woredas, with 653 households participating in the interviews. Fig. 1 reveals the diagrammatic representation of the sampling procedure followed to select the clusters and thereby households for the study. A multi stage cluster sampling was used to select clusters from each woreda, and then systematic random sampling was employed to select households within each cluster.

In addition, the data collection process included observing the presence of functional LLINs hung over the sleeping area in the household or pastoral tents.

#### Data collection and management

Data collection was conducted using hard copies of pretested and thoroughly corrected questionnaires. The principal investigator checked the accuracy, completeness, and validity of the data collected before and during the data entry process. The data entry was done by using Epidata software version 4.6.0.2. The data entry dashboard was created using “Epidata Manager” to apply skip patterns accurately and the double entry feature to reduce clerical errors. Furthermore, the researcher performed regular backups of the data to ensure its security and prevent loss in the event of technical difficulties.

### Data analysis

The data was entered into the Epidata client feature version 4.6.0.2 to reduce clerical errors and exported to SPSS version 28 for statistical analysis. The researcher cleaned the data by removing outliers and missing values before conducting descriptive and inferential analyses. Descriptive statistics computed include frequencies, measures of central tendencies, and dispersion. Independent predictors of the outcome of interest, malaria prevention practices of pastoralists of the South Omo Zone, were identified through factor analysis, chi-square, and univariate analyses.

Binary logistic regression analysis was carried out to identify the associated candidate predictors for multivariable logistic regression analysis with a significance level of less than or equal to 0.20, in addition to expert judgment and public health importance. The dependent variable with the binary outcome that was used for association testing is the practice of malaria prevention by the households [35].

Multiple variable logistic regression analysis at a significance level of ≤ 0.05 was conducted to determine the impact of various factors on the likelihood of engaging in malaria prevention behaviours. Predictors with an adjusted odds ratio of ≥ 1.5 and a P-value of ≤ 0.05 were significantly associated with practicing malaria prevention and were identified as possible determinants of engaging in malaria prevention behaviours.

#### Ethical consideration

The researcher obtained ethical clearance from the University of South Africa’s Department of Health Studies Research Ethics Committee (Reference number: 67140386_CREC_CHS_2021) and an official support letter from the former SNNPR Health Bureau, South Omo Zonal Health Department, and five woreda health offices. Informed written consent was obtained from participants, and data was anonymized for confidentiality purposes to ensure the privacy and protection of participants’ personal information. The study adhered to all necessary ethical guidelines and protocols to uphold the rights and well-being of all individuals involved in the research.

## Results

### Sociodemographic characteristics of the of the respondents

A total of 653 pastoral households were included in the face-to-face interview from 5 purposively selected high-malaria burden woredas in the South Omo Zone, South Ethiopia.

In terms of gender mix, 414 (63.4%) of the study respondents were male; the age range of the respondents was between 18 and 70 years (median of 37.0 and standard deviation of 12.62 years); 493 (75.6%) of them had no formal education; and 522 (80.0%) of them were married.

The relative socioeconomic status of the respondents in terms of the wealth index quantiles was found to be 103 (15.8%) were extremely poor, 158 (24.2%) were poor, 129 (19.8%) had middle-class incomes, 135 (20.7%) were wealthy, and 128 (19.6%) were extremely wealthy.

Three-quarters (493, or 75%) of study respondents did not attend school, while 160 (or 25%) received some form of formal education. The vast majority of those who have attended school, 104 (65%), have only completed primary school, with a smaller percentage completing secondary school or higher education.

Regarding the religious distribution of the study respondents, the majority (315, or 48.2%) of the respondents were followers of indigenous religion. The study included 9 out of 15 ethnic groups in the South Omo zone administration, with Hamer and Malie accounting for more than half.

### Awareness of the study respondents about malaria prevention and control

From 653 study respondents included in the study, 332 (50%) reported having heard or seen malaria prevention information. Health extension workers account for 267 (67%), community events for 73 (17%), and radio, posters, and television for the remainder of malaria prevention information.

Regarding the primary cause of malaria, 333 (51%) of respondents identified mosquitos as the primary cause of malaria, with contaminated water coming in second 191 (22.4%), contaminated food 81(9.5%), those who didn’t know what caused it 68 (8.1%), and close contact with an infected person (4.1%). Other less common causes include 12 (1.4%) mentions of eating sugar cane, 14 (1.6%) mentions of bad weather, 6 (0.7%) mentions of eating boiled corn, and the remaining 8 (0.9%) mentions of various other factors as the primary cause of malaria, including things like poor sanitation, stagnant water, poverty, and inadequate access to health care.

When asked about malaria prevention methods they are aware of, sleeping under nets comes in first (459, 48.4%) among 948 responses from 653 respondents, followed by draining stagnant water (190, 20%), using traditional medicine (156, 16.5%), not knowing of any methods (81, 8.5%), spraying the house with chemicals (51, 5.6%), and others (11, 1.2%).

More than half, 425 (65.1%) of the respondents were found to be knowledgeable about malaria preventative measures, while 228 (34.9%) were not knowledgeable.

The proportion of respondents with a good attitude towards malaria prevention and treatment-seeking aspects in this study was 316 (48.8%), while that with a poor attitude was 337 (51.2%).

### The access to and use of the vector control measures: long-lasting insecticide-treated nets (LLINs)

Nowadays, long-lasting insecticide-treated nets (LLINs) remain the most effective and sustainable malaria prevention tool in use [35].

Regarding the long-lasting net coverage, out of the 653 households included in this study, 393 (60.2%) reported having at least one long-lasting insecticidal net (LLIN). During the five years preceding the study, 733 LLINs were distributed to 393 households, with each household receiving at least one LLIN.

Among the 393 households with at least one LLIN, 163 had one, 134 had two, 83 had three, 12 had four, and only one had five LLINs. However, the remaining 260 (39.8%) households did not have any LLINs.

It was found that, 264 (40%) of the 653 households surveyed responded that at least one household member had slept under the LLIN the night before the survey.

Regarding retention of LLINs at a household level, data collectors observed 493 long-lasting nets out of the 733 distributed to the 393 households in the study districts, with an average of 1.25 LLINs per household. There was a discrepancy between the number of LLINs distributed and observed, with 240 LLINs unaccounted for. The rate of loss was highest among those who received two or more long-lasting nets per household. Fig 2 revels the number of long-lasting nets reported as received by households and observed during the data collection period.

The majority of LLINs, 714 (94%), were obtained from government-organized distribution campaigns, while the rest were acquired through other sources, such as NGOs, or purchased privately.

### The access to and use of the vector control measures: indoor residual spray (IRS)

Concerning the IRS application, 195 of the study’s households (29.8%) reported that their homes had been treated with insecticides in the 12 months preceding the survey. There were 382 (58.5%) households that responded their homes had not received an IRS application in the 12 months preceding the survey due to different reasons. Fig. 3 reveals the indoor residual spray (IRS) chemical application in the 12 months prior to the survey of households in the study area.

The reasons stated by 382 households for not applying to the IRS for their homes include: 259 (68%) participants mentioned that the spray team did not visit theirs; the other 60 (16%) expressed concerns about the effectiveness of the IRS, claiming that it did not effectively kill mosquitoes; and 63 (16% of participants) mentioned logistical challenges such as the lack of water to mix the chemicals or difficulties in removing household materials for the IRS application.

### Malaria prevention practice

From the total 653 respondents in this study, 221 (33.8%) reported having practised at least one method to prevent malaria in the 12 months prior to the survey. The majority of households (432, or 66.15%) in the study area did not practise an adequate combination of methods to prevent malaria. The most reported method was the use of long-lasting insecticide-treated nets, with 87 respondents (39%) stating that they used them, followed by the use of indoor residual insecticide spray, reported by 42 respondents (19%), and the clearing of stagnant water sources, mentioned by 22 respondents (10%); the use of traditional medicine was reported by 29 respondents (13%). Other methods mentioned included the use of mosquito repellents, mentioned by 10 respondents (4%), and other methods such as taking antimalaria medicines and wearing protective clothing.

However, there were respondents who stated they had been doing nothing in the past 12 months prior to the survey to prevent malaria, accounting for 40 (18%) of the total respondents.

### Result of the logistic regression in relation to malaria prevention practice

In a multivariable logistic regression, it was revealed that the odds of practicing at least two and possibly more malaria prevention measures were significantly higher in the wealth index quantiles of middle-income (AOR = 2.746 (1.534–4.915)) and wealthier (AOR = 2.608 (1.461–4.655)) relative wealth distribution households as compared to poorer households and women-headed households (AOR = 1.616 (1.115–2.334)) as compared to men-headed households. Table 1 displays the summary of multivariable logistic regression analysis, which shows the strength of the association between sociodemographic predictors and respondents’ use of malaria prevention.

**Table 1:** Logistic regression analysis showing the strength of association between the sociodemographic predictors and the practice of malaria prevention by respondents (N=653)

| Sr no | VARIABLES |  | FREQUENCY | CRUDE OR* (95% CI**) | P_VALUE | ADJUSTED OR (95% CI) |
| --- | --- | --- | --- | --- | --- | --- |
| 1 | Sex | Male | 414 | 1 | 0.011 | 1 |
|  |  | Female | 239 | 1.508(1.081-2.104) |  | 1.616(1.115-2.334) *** |
| 2 | Age (years) | Young adult (18-35) | 292 | 2.17(1.26-3.72) | 0.125 | 1.583(0.880 2.846) |
|  |  | Middle aged adult (36-55) | 270 | 1.53(0.88-2.66) | 0.387 | 1.293(0.722-2.317) |
| | | Elder ($\geq 56$ years) | 91 | 1 | | 1 |
| 3 | Education | No education | 493 | 1 |  | 1 |
|  |  | Primary school (grade1-8) | 104 | 1.204(0.772-1.878) | 0.471 | 1.194(0.738-1.932) |
|  |  | Secondary (grade 9-12) | 47 | 2.090(1.144-3.818) | 0.262 | 1.453(0.756-2.792) |
|  |  | Higher (College & above) | 9 | 4.361(1.077-17.666) | 0.092 | 3.472(0.815-14.783) |
| 4 | Marital status | Married | 528 | 0.809(0.13-4.88) | 0.881 | 0.867(0.134-5.602) |
|  |  | Never married | 35 | 1.00(0.14-6.77) | 0.904 | 0.884(0.120-6.510) |
|  |  | Widowed | 72 | 0.362(0.05-2.38) | 0.541 | 0.545(0.078-3.821) |
|  |  | Divorced | 12 | 1.500(0.18-12.45) | 0.783 | 1.363(0.150-12.351) |
|  |  | Separated | 1 | 0.000 | 1.000 | 0.000(0.000-0.000) |
|  |  | Living with a/man or woman | 5 | 1 |  | 1 |
| 5 | Wealth index<br>(quantiles) | Extremely poor | 122 | 1 |  | 1 |
|  |  | Poor | 98 | 0.921(0.513-1.651) | 0.580 | 0.845(0.465-1.535) |
|  |  | Middle -income | 78 | 3.169(1.797-5.589) | 0.001 | 2.746(1.534-4.915) *** |
|  |  | Wealthier | 70 | 2.897(1.650-5.087) | 0.001 | 2.608(1.461-4.655) *** |
|  |  | Wealthiest | 64 | 0.955(0.520-1.755) | 0.900 | 0.961(0.514-1.794) |

Legend for tables: \*OR- odds ratio, \*\* CI- confidence interval \*\*\* Statistically significant association by adjusted OR

Apart from sociodemographic factors, malaria prevention knowledge was also significantly associated with practicing malaria prevention at the household level. Table 2 displays the multivariable logistic regression analysis, which shows the strength of the association between malaria prevention aspects and respondents’ use of malaria prevention. Being knowledgeable about malaria prevention aspects outperformed not being knowledgeable (AOR = 3.399 (2.106–5.486)), and, in a similar way, having the long-lasting insecticide-treated nets at home (AOR = 7.881 (2.309–26.896)) as compared to not having them at the times of the study.

**Table 2:** Logistic regression analysis showing the potential malaria programme related predictors on influencing practice of malaria prevention by respondents (N=653)

| SR<br>NO | VARIABLES |  | FREQUENCY | CRUDE OR* (95% CI**) | P_VALUE | ADJUSTED OR (95% CI) |
| --- | --- | --- | --- | --- | --- | --- |
| 1. | Heard message about malaria in the last 6 months? | Yes | 321 | 2.198(1.577-3.063) | 0.115 | 1.368 (0.926-2.020) |
|  |  | No | 332 | 1 |  | 1 |
| 2. | Availability of LLINs | Yes | 393 | 13.41(8.07-22.27) | 0.001 | 7.881(2.309-26.896)<br>*** |
|  |  | No | 260 | 1 |  | 1 |
| 3. | Number of LLINs | No LLIN | 260 | 1 |  | 1 |
|  |  | 1 LLIN | 163 | 9.547(5.450-16.725) | 0.994 | 0.996(0.308-3.220) |
|  |  | 2 LLINs | 134 | 13.465 (7.562-23.976) | 0.749 | 1.213(0.373-3.946) |
|  |  | 3 LLINs | 83 | 27.808(14.398-53.707) | 0.134 | 2.547(0.749-8.663) |
|  |  | ≥ 4 LLINs | 13 | 10.872(3.320-35.603) | 0.994 |  |
| 4. | Knowledge of individuals | Knowledgeable | 428 | 5.65(3.66-5.8.72) | 0.000 | 3.399(2.106-5.486)<br>*** |
|  |  | Not Knowledgeable | 228 | 1 |  | 1 |
| 5. | Attitude of individuals | Favourable attitude | 316 | 1.643 (1.185-2.2.78) | 0.680 | 1.085(0.736-1.599) |
|  |  | Unfavourable attitude | 337 | 1 |  | 1 |

## Discussion

In this study, 282 (67%) of respondents said they got their malaria information from health extension workers, followed by 73 (17%) who said they got it from community events, 41 from television, and 15 from posters and radio. Similarly, in a recent study conducted by Menjetta in 2019 in the Halaba zone in Southern Ethiopia, 370 respondents (87.9%) reported getting the information from health extension workers, while about 30 (7.9%) said they got it from the media, and In a study conducted by Aragie in the West Belessa district of north Ethiopia in 2020, 369 (48.1%) respondents got information from health workers, 185 (24.1%) from relatives, and 165 (21.5%) from community events [36, 37].

In the pastoral communities of the South Omo Zone in Ethiopia, this suggests that health extension workers are the most effective source of malaria health information, followed by community malaria events, as they were in the West Belessa and Halab districts. However, it is important to note that the effectiveness of these sources may vary depending on the cultural and social context of the community.

Regarding awareness about the causes of malaria in this study, nearly more than half (433, or 51%) of the responses from multiple categories categorized as “mosquitoes” were followed by 191 (22.4%) for contaminated water, 69 (8.1%) for contaminated food, and 35 (4.1%) for close contact with a malaria-infected person.

However, in a recent study by Menjetta [37] in Halaba Zone in Ethiopia, those who responded as mosquitoes were found to be 267 (63.4%) of the study respondents who thought that mosquitoes were responsible for the transmission, and in another study done among nomads in Chad by Aliyo, Golicha [38], 68.3% of respondents responded as malaria mosquito bites.

The reason for this discrepancy between the two sets of results may be due to the socio-cultural, structural, socio-economic, and access to health services differences in the Halaba community, which is sedentary and urbanised and has had more exposure to health education programmes and interventions as compared to the pastoralist communities in the South Omo zone.

According to a study conducted in South Gondar, Amara Region, Ethiopia, by Flatie and Munshea [39], 44 (11.3%) of the participants did not know the cause of malaria; similarly, in this study, 81 (9.5%) of the heads of those surveyed responded that they didn’t know what causes malaria.

This shows that a significant number of respondents’ lack knowledge about malaria’s cause and have misconceptions about other causes, highlighting the need for health education and awareness activities in nomadic and seminomadic pastoral communities.

This study revealed the respondents’ beliefs about methods that can prevent malaria as follows: sleeping in long-lasting nets (459; 48.4%); draining stagnant water (190; 29.0%); using traditional methods like herbs and other medicinal practices like cleaning the house (156; 16.5%); not knowing (81; 8.5%); IRS 51 (5.4%); and other methods like eating good food, drinking clean water, and avoiding contact with infected persons (11; 1.2%).

From these findings, it can be speculated that the majority of respondents believe that sleeping in long-lasting nets and draining stagnant water are the most effective method for preventing malaria. However, it is also important to note that a significant portion of respondents also believe in using traditional methods and draining stagnant water, which suggests a need for culturally sensitive approaches to malaria prevention.

In this study, 651 (65.1%) respondents were categorised as knowledgeable, while 228 (34.9%) were unknowledgeable about malaria prevention and treatment related information.

While Studies in South Gondar, North-western Ethiopia, Cameroon, and Mozambique found that the respondents’ knowledge of malaria prevention was higher than it was in this study[39–41].

The differences in this study could be attributed to a variety of factors, including differences in education levels, malaria programme-related activities, access to healthcare services, lifestyles, cultural beliefs, and geographical location.

It is important to consider these factors when designing and implementing interventions to control and prevent malaria, as they can significantly impact the effectiveness of such interventions in settings like nomadic pastoral communities.

The current study assessed participants’ attitudes towards malaria prevention and treatment seeking. It was found that 319 (48.8%) of participants had positive attitudes towards these aspects, while 334 (51.2%) had negative attitudes.

This figure was also found to be lower than the attitude evaluation results obtained from studies conducted in South Gondar, North-western Ethiopia, and Limpopo Province, South Africa, by [37, 42].

In terms of ownership of the long-lasting insecticide-treated nets, the long-lasting net availability at the household level is 60.2%, which is slightly higher than that of Mogadishu and the Borena zone in Ethiopia.

According to a study conducted in Mogadishu, Somalia, the proportion of households that owned LLINs was 37.9%, and another study by Aliyo, Golicha [38] among pastoral households in the Borena zone in Ethiopia found it to be 47%.

Similarly, this study found that (40%) of the households surveyed said at least one member had slept under the LLIN the night before the survey.

This indicates a moderate level of LLIN usage difference can be attributed to varying cultural beliefs, practices, and access to LLINs among different populations

In this study, of the 336 households with at least one long-lasting net, 264 (78.6%) of the households reported having used it the night before the survey. Of the 230 households that owned two or more long-lasting nets, 93 (40.8%) of the households reported using them, and 139 (60.6%) did not, and the results get worse with households with more than two nets.

According to a study conducted by Hambisa, Debela [43], 90% of households had long-lasting nets, and 68.3% were using the nets the night before the survey. Like this, the national malaria indicator survey carried out in Ethiopia in 2015 showed that 61 percent of the population slept under an LLIN the night before the survey, and the percentage was higher for those with one LLIN.

These two studies’ findings confirm that households with a single LLIN use more long-lasting nets than households with multiple LLINs.

This means that providing one LLIN per household and multiple LLINs needs to be re-evaluated as a strategy for malaria prevention. It may be more effective to focus on distributing and promoting the appropriate use of long-lasting nets to households with no or insufficient LLINs.

However, the prevalence of using LLINs was found to be lower among the nomadic pastoralists of South Omo in this study compared to the national survey, possibly due to the frequent displacement of pastoralists, the limited availability of LLINs in remote areas, and a lack of knowledge about the value of using long-lasting nets.

Generally, the use of long-lasting nets among the pastoral community of the South Omo was found to be relatively low, despite the high malaria prevalence in the area; the possible explanations could be inadequate access to LLINs due to their geographical remoteness, a lack of awareness of the health benefits associated with their use, as well as socio-cultural issues, such as their nomadic lifestyle. In addition, the study respondents who didn’t use the long-lasting nets they had given the following reasons: the nets being too hot and uncomfortable, the colour not being their preference, the fact that they did not kill mosquitoes, the extra savings for future use, the lack of mosquitoes and malaria, and not liking the smell of long-lasting nets.

Indoor residual spray (IRS) is recommended by The World Health Organization (WHO) as an effective malaria control method in high- and medium-risk areas [44]. The WHO has recommended indoor residual spray (IRS) as a key strategy for malaria prevention, control, and elimination with a high return on investment when used in conjunction with LLINs and improved diagnostic testing and treatment[45]. If implemented with sufficient political commitment and social acceptance of the IRS, adequate program and health system capacity to deliver high-quality, timely care, and adequate knowledge and understanding of the communities served, the IRS is likely to have a major impact on malaria transmission in many settings [44].

According to the study conducted byMekasha, Daba [46], in the Shewa Robit district of Ethiopia, the indoor residual spray coverage at the household level was 56.5%. In another study conducted by Dimas, Sambo [47] in Nasarawa State, North-Central Nigeria, 99.3% replied their houses were sprayed with IRS chemical.

In this study, however, the IRS coverage was found to be in 194 (or 29.8%) of the pastoral households of the South Omo Zone in Ethiopia, which reported that their indoor residual spray coverage had been carried out during the previous 12 months’ indoor residual spray coverage. From this, it can be concluded that IRS coverage in pastoral households is much lower than in other areas of the country.

The main reasons for this low coverage can be attributed to a lack of access to the necessary resources, poor communication, the limited access of pastoral households to IRS programs, a lack of awareness and the scarcity of the IRS chemical spray budget, the regular camp shifts to search for pasture by the pastoral communities, and their nomadic lifestyle.

In a community-based study conducted in the West Bellessa district of the Amhara region by Aragie and Wolde [48], 21.1 percent of the rural community there used long-lasting insecticide nets (LLINs) to prevent malaria and 80.5% used insecticide residual spray (IRS), for a combined average of 50 percent by both methods.

In this study, 185 households (28.3%) used no method of malaria prevention, while 247 households (30.7%) used a single method of malaria prevention, which is not adequate to achieve malaria prevention in high transmission settings like the one in which this study took place. While 187 households (28.5%) used two methods of malaria prevention, 34 households (5.2%) used three or more methods of malaria prevention.

This result suggests the need for more education and awareness campaigns about the significance of using multiple methods of malaria prevention in high transmission settings like the nomadic pastoral communities of South Omo, Ethiopia, because relying solely on one method or not using any method may not be effective. To achieve better malaria prevention results, it is critical to encourage the use of a variety of strategies, including indoor residual spraying, insecticide-treated bed nets, environmental related vector control and prompt treatment of cases of malaria.

According to a study conducted byAragie and Wolde [48], the odds of practicing malaria prevention are lower in the poorest, middle-class, and wealthy wealth quintiles, the female gender, in a certain village of residence, among illiterate groups, and among those with poor malaria prevention knowledge in contrast to other categories.

This study revealed that socioeconomic factors, illiteracy, and knowledge about malaria prevention are important for understanding differences in the practice of malaria prevention, except being the reverse for gender. In this study, as displayed in table 2, in multivariable logistic regression, which was used to investigate the factors associated with malaria prevention practices, the following predictors of the practice were found to have significant odds ratios: Women-headed households (AOR = 1.616 (1.115-2.334)) had significantly higher odds of using two or more malaria prevention strategies than men-headed households in the wealth index quantiles of middle-income (AOR = 2.746 (1.534-4.915)) and wealthy (AOR = 2.608 (1.461-4.655)) relative wealth distribution households as compared to poorer households and those with knowledge of malaria prevention strategies.

Furthermore, as displayed in table 2, in relation to malaria program-related factors, those with knowledge of malaria prevention strategies outperformed those without it (AOR = 3.399 (2.106– 5.486)), and those having the long-lasting insecticide-treated nets at home at the time of the survey performed better than those not having them (AOR = 7.881 (2.309–26.896)).

In contrast to previous findings by Aragie and Wolde (2021), who discovered that the odds of practising malaria prevention were lower in the poorest, middle-class, and wealthy wealth quintiles, the female gender, in a specific village of residence, among illiterate groups, and among those with poor malaria prevention knowledge, this study discovered that the odds of practising malaria prevention were higher among the wealthy, middle-class, females having, and those with poor malaria prevention knowledge. This was an interesting finding, as it showed that even among traditionally disadvantaged populations like pastoralists, such as women and poorer households, they still had higher odds of practicing malaria prevention methods when compared to male-headed households and the possible explanations for this could include the fact that women may be more likely to prioritize their families’ health due to their decreased mobility compared to men in a pastoral setup.

Across cultures and eras, women have traditionally served as the primary carers for family health needs, ensuring the wellbeing of children, parents, and other family members [49].

The study found that factors like gender roles, education, malaria prevention knowledge, access to nets, and economic status influence households’ decision-making. So that malaria prevention strategies can be implemented in a setting like that of nomadic pastoral communities, they should focus on increasing access to preventive products and ensuring vulnerable households have the necessary knowledge and resources.

## Limitations of the study

This study is a cross-sectional study on pastoral communities in Ethiopia’s South Omo Zone. Although the findings may not be generalizable due to differences in cultural practices and environmental conditions among pastoral communities in other regions, they can help guide similar interventions in other areas by providing tailored solutions for pastoralist communities in Ethiopia’s South Omo Region.

## Conclusions

This study identified factors associated with malaria prevention and treatment practises. The study demonstrated that greater wealth, education, access to long-lasting insecticide-treated nets, and understanding of the concept of malaria prevention are all associated with increased odds of practising malaria prevention and treatment and established this association using quantitative methods.

## Data Availability

The data utilized in the research was submitted as zuped or compressed supporting information along with the manuscript. For further information, the data can be obtained upon request by contacting the corresponding author via email at.

## Acknowledgment

We extend our deepest gratitude to the Department of Health Sciences at the University of South Africa for granting me access to the tools required to carry out this study and to the pastoral community of the South Omo Zone for their willingness to participate and share their valuable insights, which were crucial in shaping the findings of this research.

We are also grateful to Mr.Tadiwos Toyota, Mr.Yohannes Negash, and Mr. Mintesnot Gujo for supervising data collection and providing hospitality during fieldwork at the South Omo zonal health department. In addition, we would like to thank my former coworkers from the Regional Health Bureau of the Southern Nations and Nationalities, as well as Mr. Misganu Endrias, Mr. Mena Mekuria, and Mr. Male Mate, for their help and support during the research.

---

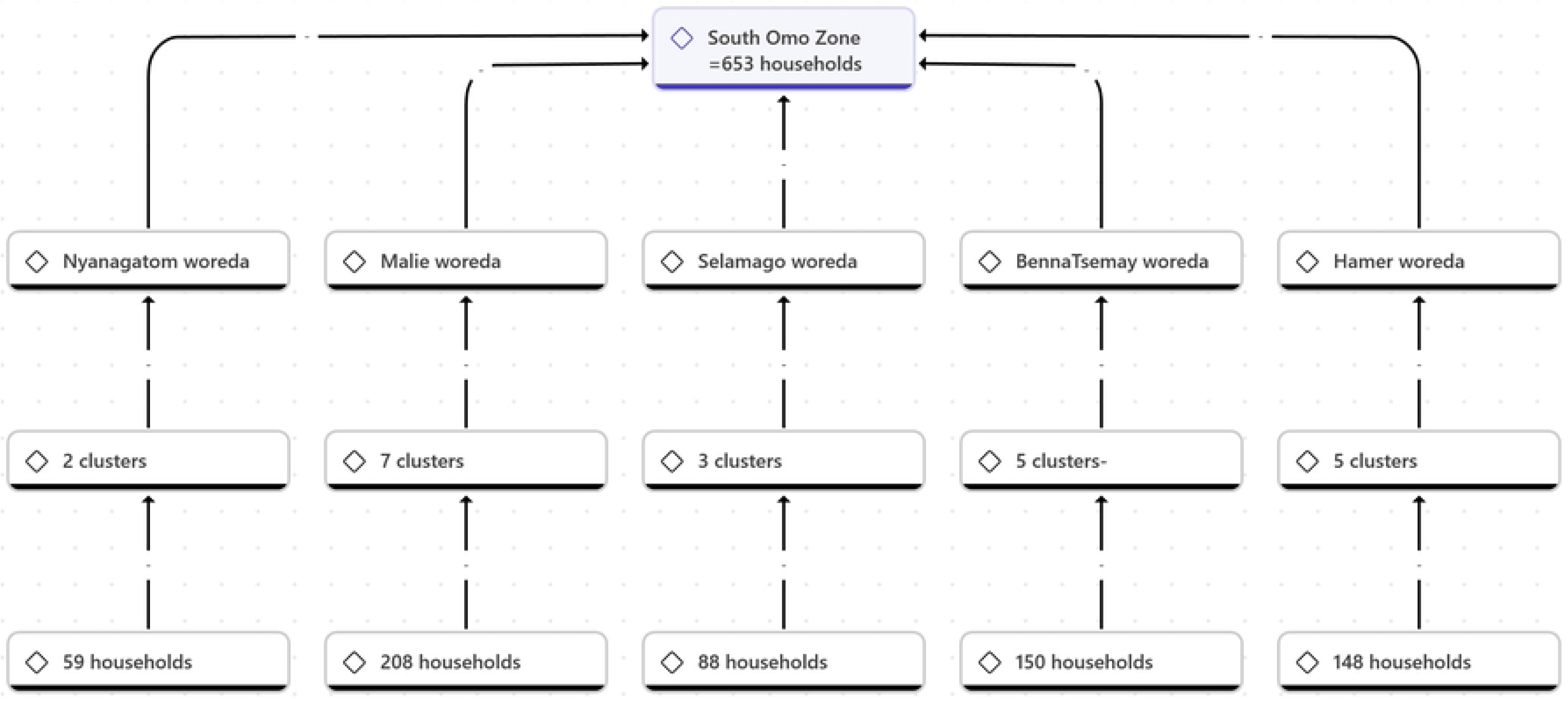

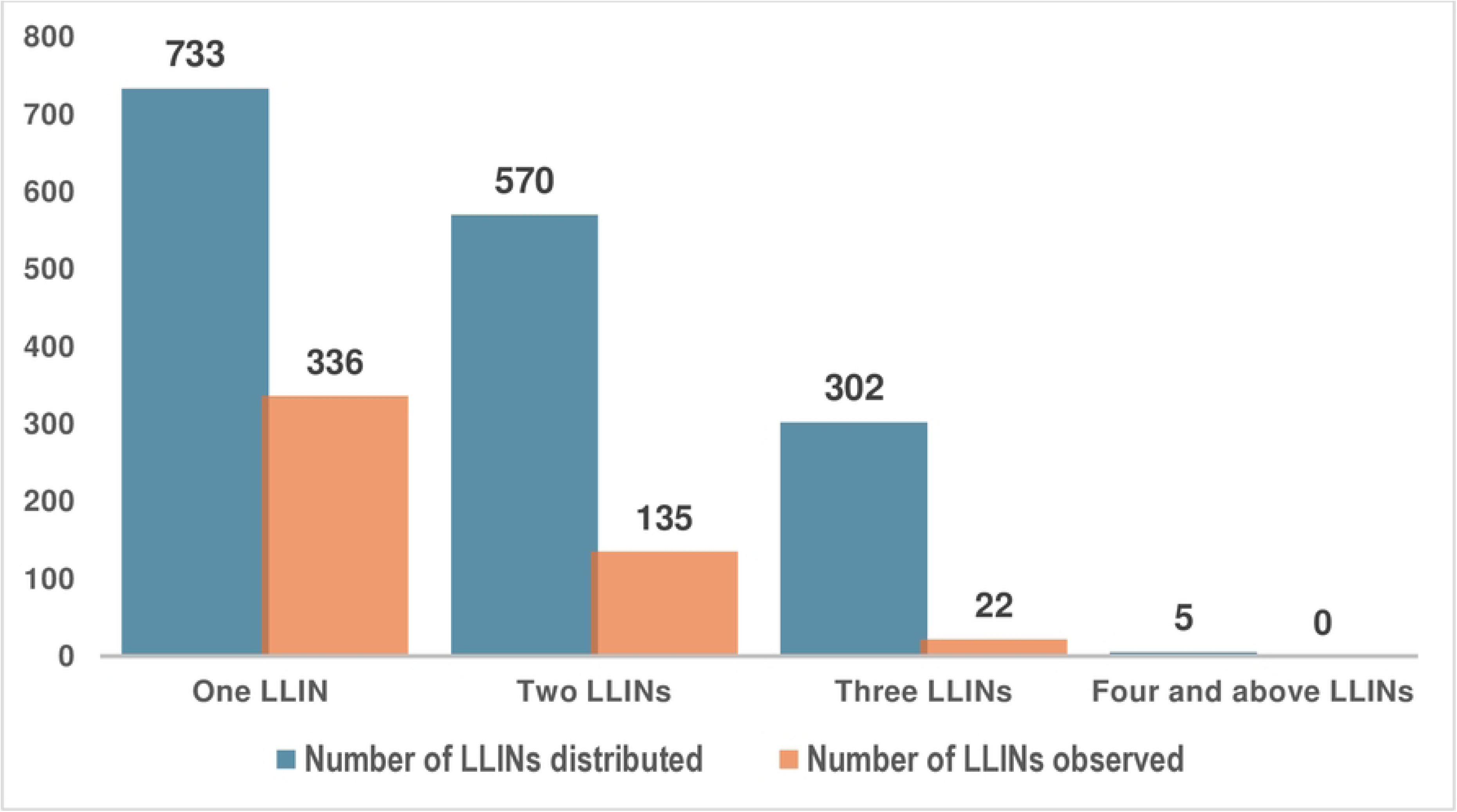

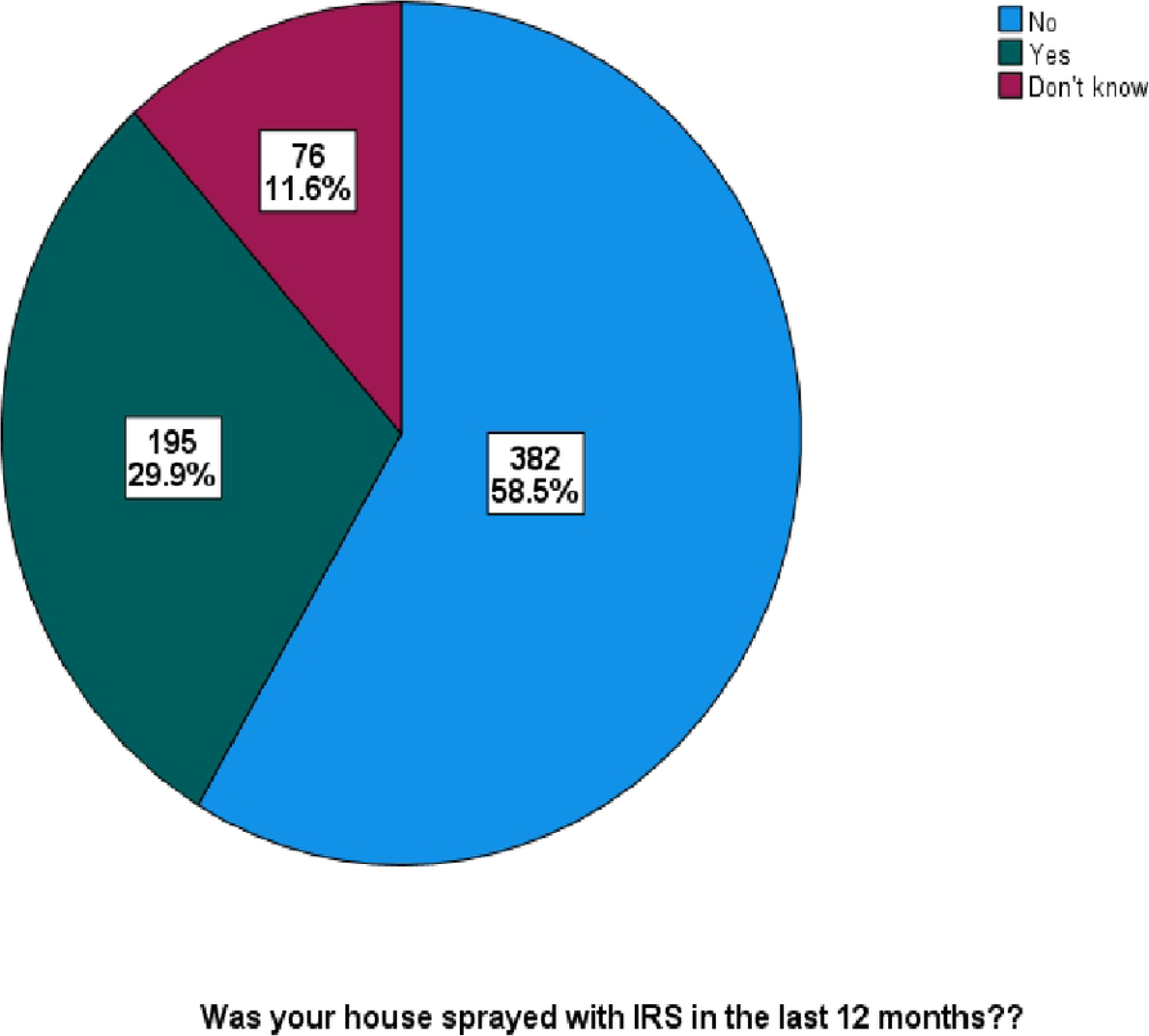

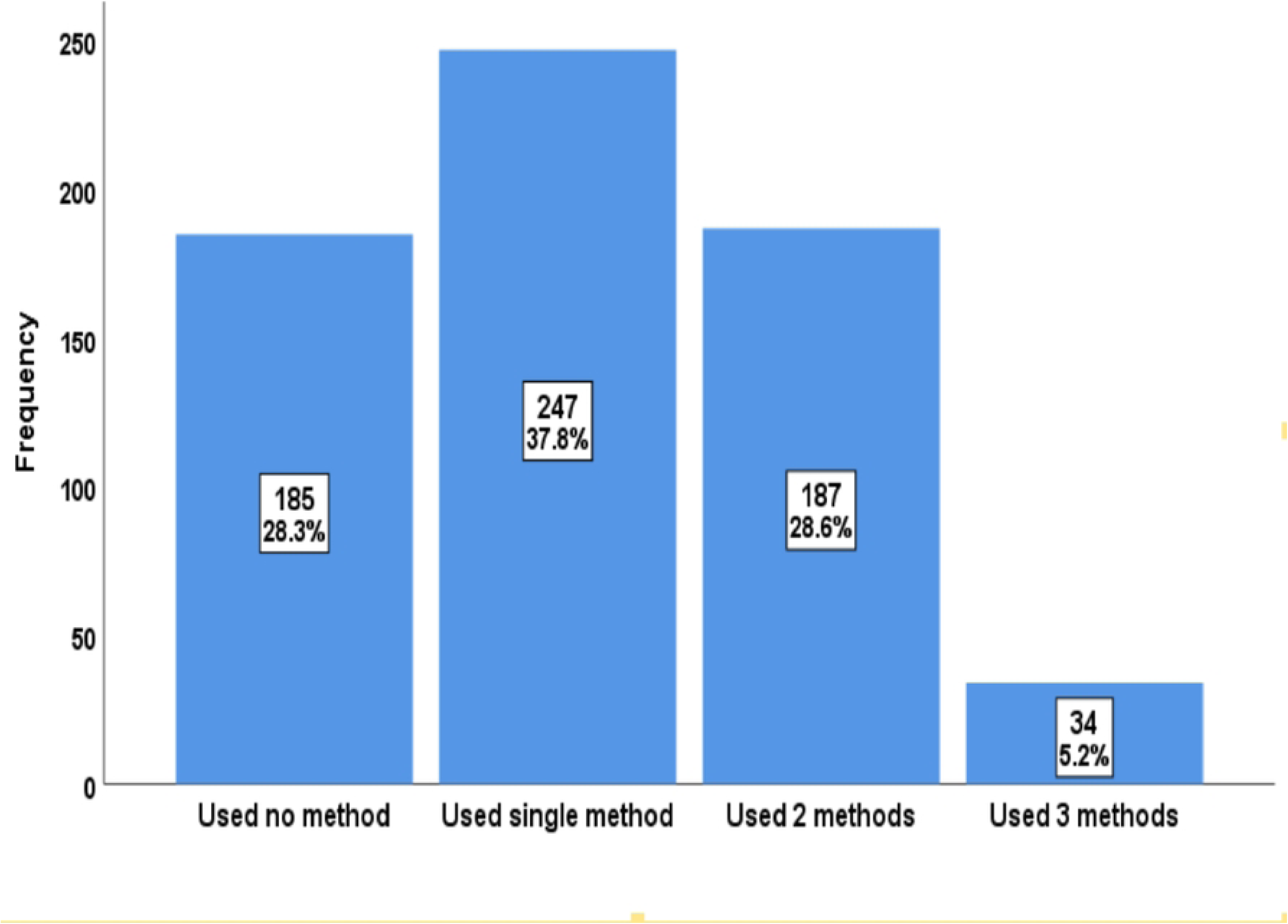

